# One-Year Safety and Effectiveness of the ISAR SUMMIT Polymer-Free Everolimus-Eluting Stent in Real-World Clinical Practice

**DOI:** 10.64898/2026.07.16.26358282

**Authors:** Praveen Chandra, Yash Paul Sharma, Rajneesh Kapoor, Rahul Singhal, Pushpraj Patel, Anupam Jena, Dileep Kumar Tiwari, Rohit Mody, Amjad Ali, Aditya Kapoor, Preeti Sharma, Viveka Kumar, Kamal Sharma, Vikas Chopra, Milind Nivrutti Kharche, Vikas Kataria, Sameer Dani, Deepak Davidson, Rajiv Agarwal, PLN Kapardy, Ripen Gupta, Rohan Ainchwar, Ashwani Mehta, Aziz Khan, Jaspal Arneja, Adnan Kastrati

## Abstract

**Aims:** Polymer-free drug-eluting stents were developed to enhance vascular biocompatibility and safety while maintaining antirestenotic efficacy. The TRANSEVER registry evaluated 12-month clinical outcomes of the polymer-free everolimus-eluting ISAR SUMMIT stent in a large, real-world population undergoing percutaneous coronary intervention.

**Methods:** This prospective, multicentre study enrolled patients with coronary artery disease undergoing PCI with the ISAR SUMMIT stent across 33 centres in India. The primary endpoint was target-lesion failure (TLF) at 12 months, a composite of cardiac death, target vessel myocardial infarction, or clinically driven target lesion revascularisation. Secondary endpoints included the patient-oriented composite endpoint (POCE) of all-cause death, any myocardial infarction, stroke, revascularization, and definite/probable stent thrombosis.

**Results:** A total of 1,000 patients were enrolled, of whom 996 completed 12-month follow-up. The cohort presented with a high-risk profile, including an acute coronary syndrome (ACS) in 89.8% of the cases and diabetes mellitus in 44.4% of them. Procedural outcomes were excellent in terms of device success and final TIMI 3 flow (achieved in all treated lesions). At 12 months, TLF occurred in 15 patients (1.5%). Definite or probable stent thrombosis was observed in 8 patients (0.8%). POCE was observed in only 21 patients (2.1%).

**Conclusions:** In this large, contemporary real-world population with a very high proportion of patients presenting with ACS, the polymer-free everolimus-eluting ISAR SUMMIT stent demonstrated favourable 12-month clinical outcomes, with low rates of target lesion failure and stent thrombosis. These results suggest that this novel device is both safe and effective for routine clinical use.

## INTRODUCTION

Drug-eluting stents (DES) have significantly improved the results of treatment of patients with coronary artery disease (CAD) by reducing the need for reintervention following percutaneous coronary intervention (PCI)^1^. Successive iterations of DES have focused on improving biocompatibility and long-term safety, particularly in response to concerns regarding delayed vascular healing and late thrombotic events associated with earlier polymer-based platforms^2^.

Two key design strategies, reduction in strut thickness and optimisation of the drug delivery, have emerged in contemporary DES development^3,4^. Thinner struts are associated with less vessel injury^5^ and improved hemodynamics, while polymer-free drug delivery has been proposed to minimise chronic inflammatory responses attributed to durable or slowly degrading polymers^6^. Parallel to these structural refinements, everolimus has become one of the most extensively studied antiproliferative agents in DES technology, owing to its predictable elution characteristics, potent inhibition of neointimal hyperplasia, and favourable safety profile^7^.

The ISAR SUMMIT is a third-generation polymer-free everolimus-eluting stent designed to enable controlled drug delivery via a microporous surface without a polymer carrier, thereby improving vascular healing and preventing restenosis. Given the distinct clinical profile of patients undergoing PCI in India, characterised by a high prevalence of diabetes and acute coronary syndromes (ACS), real-world evidence is needed to evaluate the performance of ISAR SUMMIT in this setting. In this study, we present the final 12-month outcomes of 1,000 enrolled patients in the TRANSEVER registry, providing a comprehensive assessment of the safety and effectiveness of the ISAR SUMMIT stent in real-world practice.

## METHODS

### Study design and population

The TRANSEVER registry was a prospective, multicenter, real-world study evaluating the polymer-free everolimus-eluting ISAR SUMMIT stent in patients with CAD undergoing PCI. The detailed study design and conduct have been previously reported^8^.

Briefly, adult patients (≥18 years) undergoing PCI with the study device were enrolled across 33 centres in India from August 2022 onward. The multicenter design ensured broad geographic representation and diverse clinical practice patterns, thereby enhancing the generalisability of the findings to real-world settings.

Procedural strategy, access site, lesion preparation, and use of adjunctive intravascular imaging were left to the discretion of the operator. Dual antiplatelet therapy was prescribed according to contemporary guideline recommendations.

Written informed consent was obtained from all patients prior to enrolment. The study was approved by the institutional ethics committees (IEC) of all participating centres and registered with the Clinical Trials Registry of India (CTRI/2022/07/044472). An independent Data Safety Monitoring Board (DSMB) oversaw the conduct of the study, and the clinical adverse events were adjudicated according to predefined definitions.

### Endpoints

Clinical follow-up was planned at 1 month, 6 months, and 12 months after the index procedure. Follow-up assessments were performed through on-site outpatient visits or telephone contact, as applicable. The primary endpoint was target-lesion failure (TLF) at 12 months, a composite of cardiac death, target vessel myocardial infarction (TV-MI), or clinically driven target lesion revascularisation (TLR). Secondary endpoints included the patient-oriented composite endpoint, a composite of all-cause mortality, any myocardial infarction, stroke, and repeat revascularisation at 12 months. Device success was also assessed and defined as successful delivery and deployment of the study stent at the intended target lesion with final TIMI 3 flow and without device-related complications. Stent thrombosis was assessed as a safety endpoint and classified as definite or probable according to Academic Research Consortium (ARC) criteria.

### Statistical analysis

Statistical analyses were performed using SAS 9.1, R 3.1 and Python 3.2. Continuous variables are presented as mean ± standard deviation, and categorical variables as counts and percentages. Time-to-event outcomes were analysed using the Kaplan–Meier method. Patients were followed-up for 12 months or until the time of last contact or consent withdrawal for those who were not able to complete the planned 1-year follow-up.

## RESULTS

### Patient Demographics and Clinical Characteristics

A total of 1,000 patients were enrolled in this study. The mean age of the patients was 60.5 years, and 22.2% of them were women. Diabetes mellitus was present in 44.4% of the patients, and 89.8% of them presented with ACS (Table 1).

**Table 1.** Baseline clinical characteristics.

| **Characteristic** | **Patients**<br>**n = 1,000** |
| --- | --- |
| **Age, (years)** | 60.5 ± 11.0 |
| **Female sex** | 222 (22.2) |
| **Diabetes mellitus** | 444 (44.4) |
| **Hypertension** | 603 (60.3) |
| **Dyslipidaemia** | 613 (61.3) |
| **Prior myocardial infarction** | 134 (13.4) |
| **Prior PCI** | 107 (10.7) |
| **Acute coronary syndrome** | 898 (89.8) |
| **Unstable Angina** | 450 (45.0) |
| **STEMI** | 264 (26.4) |
| **NSTEMI** | 184 (18.4) |
| **Multivessel disease** | 604 (60.4) |

*Data are presented as mean ± SD or n (%).*

### Angiographic and procedural characteristics

The study population included a high proportion of patients with multivessel disease and complex lesions. Among 1,628 lesions treated, 1480 ISAR-SUMMIT stents were implanted. The most frequently lesion location was in the left-anterior descending artery (LAD, 43.6%), followed by the right coronary artery (RCA, 30.7%). Device success was assessed at the stent level and attained in 99.5% cases (Table 2).

**Table 2.**
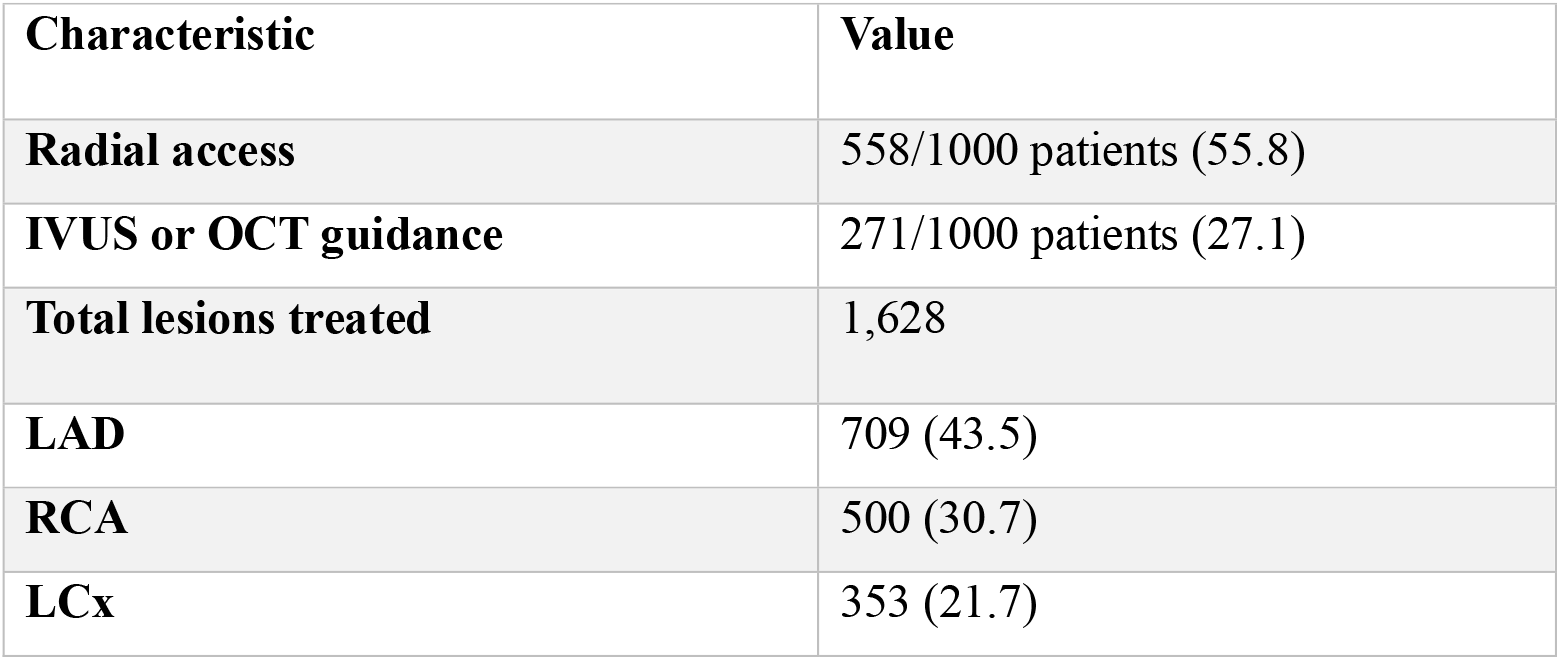
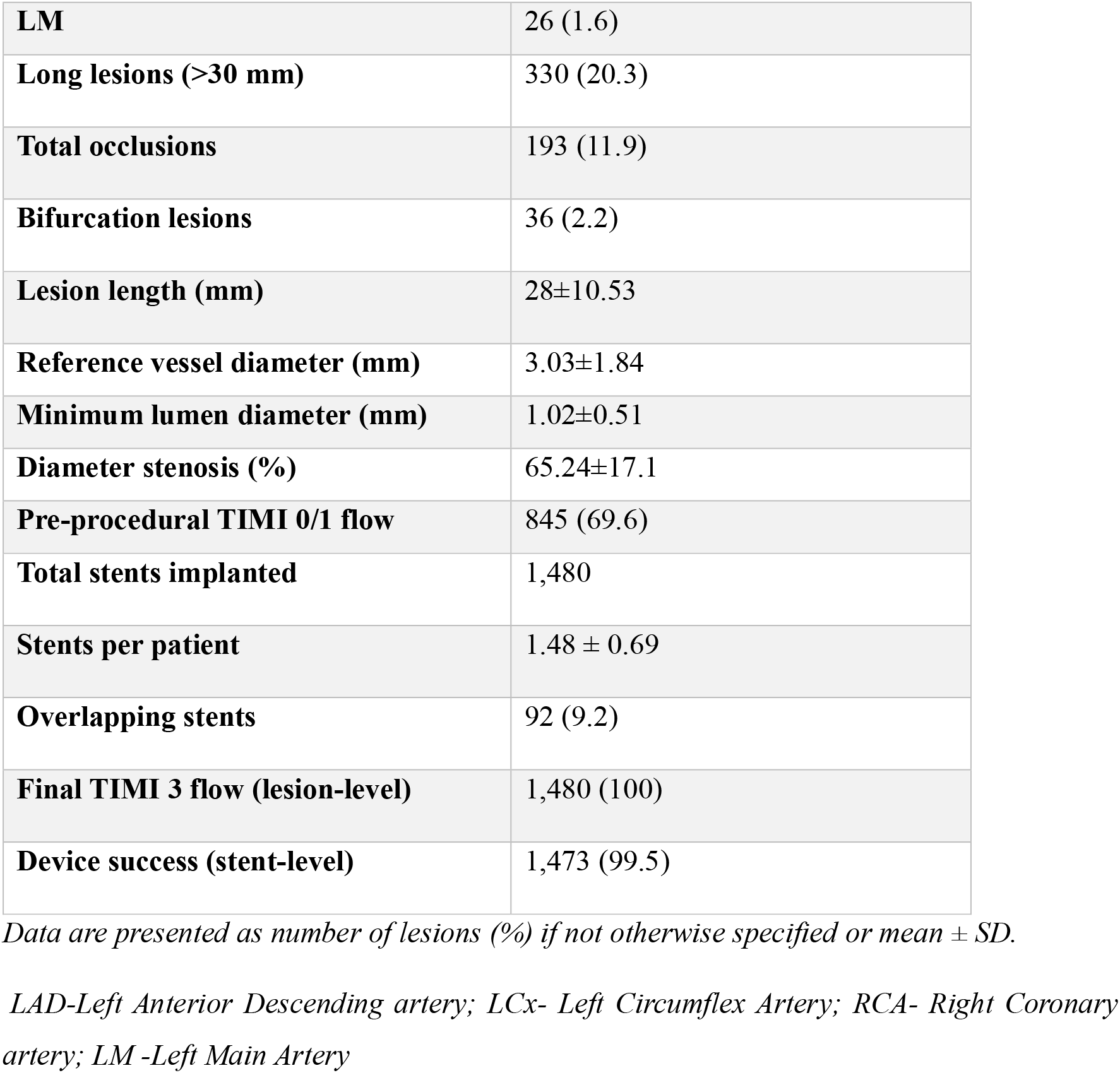
Angiographic and procedural characteristics.

### Clinical outcomes at 12 months

A 12-month clinical follow-up was available for 996 patients; two patients were lost to follow-up, and two withdrew consent. At 12 months, the primary endpoint of TLF occurred in 15 patients (1.5%). These patients experienced a total of 25 component events, comprising 10 cardiac deaths, 10 target vessel myocardial infarctions, and 5 clinically driven target lesion revascularisations. Definite or probable stent thrombosis was observed in 8 patients (0.8%).

Even when considering more broadly the adverse events as reflected by the POCE, it is evident that patients treated with the ISAR-SUMMIT stent have an excellent overall prognosis, with 21 of them (2.1%) experiencing a POCE during the follow-up period (Table 3).

**Table 3.** Clinical outcomes at 12 months.

| **Endpoint** | **Patients,**<br>**n = 1,000** |
| --- | --- |
| **Primary endpoint** |  |
| **Target-lesion failure** | 15 (1.5) |
| **Individual components of target-lesion failure** |  |
| **Cardiac death** | 10 (1.0) |
| **Target-vessel myocardial infarction** | 10 (1.0) |
| **Target-lesion revascularisation** | 5 (0.5) |
| **Secondary endpoints:** |  |
| **Patient-oriented composite endpoint (POCE)\*\*** | 21 (2.1) |
| **Individual components of POCE** |  |
| **All-cause death** | 13 (1.3) |
| **Any myocardial infarction** | 13 (1.3) |
| **Stroke** | 1 (0.1) |
| **Repeat revascularisation** | 8 (0.8) |
| **Stent Thrombosis** |  |
| **Definite** | 6 (0.6%) |
| **Probable** | 2 (0.2%) |

*Data are presented as n (%)*

The Kaplan-Meier analysis over 12 months demonstrates a low TLF incidence (Figure 1) and definite or probable stent thrombosis (Figure 2), reflecting the favourable 1-year safety and efficacy profile of the ISAR SUMMIT stent. The event curves show that most of the events were observed within the first 6 months; there after, there was a plateau with a very low number of events occurring between 6 and 12 months. This temporal pattern indicates early event accrual with sustained stability thereafter, reflecting consistent performance of this stent over time (Figs. 1 and 2).

**Figure 1.**
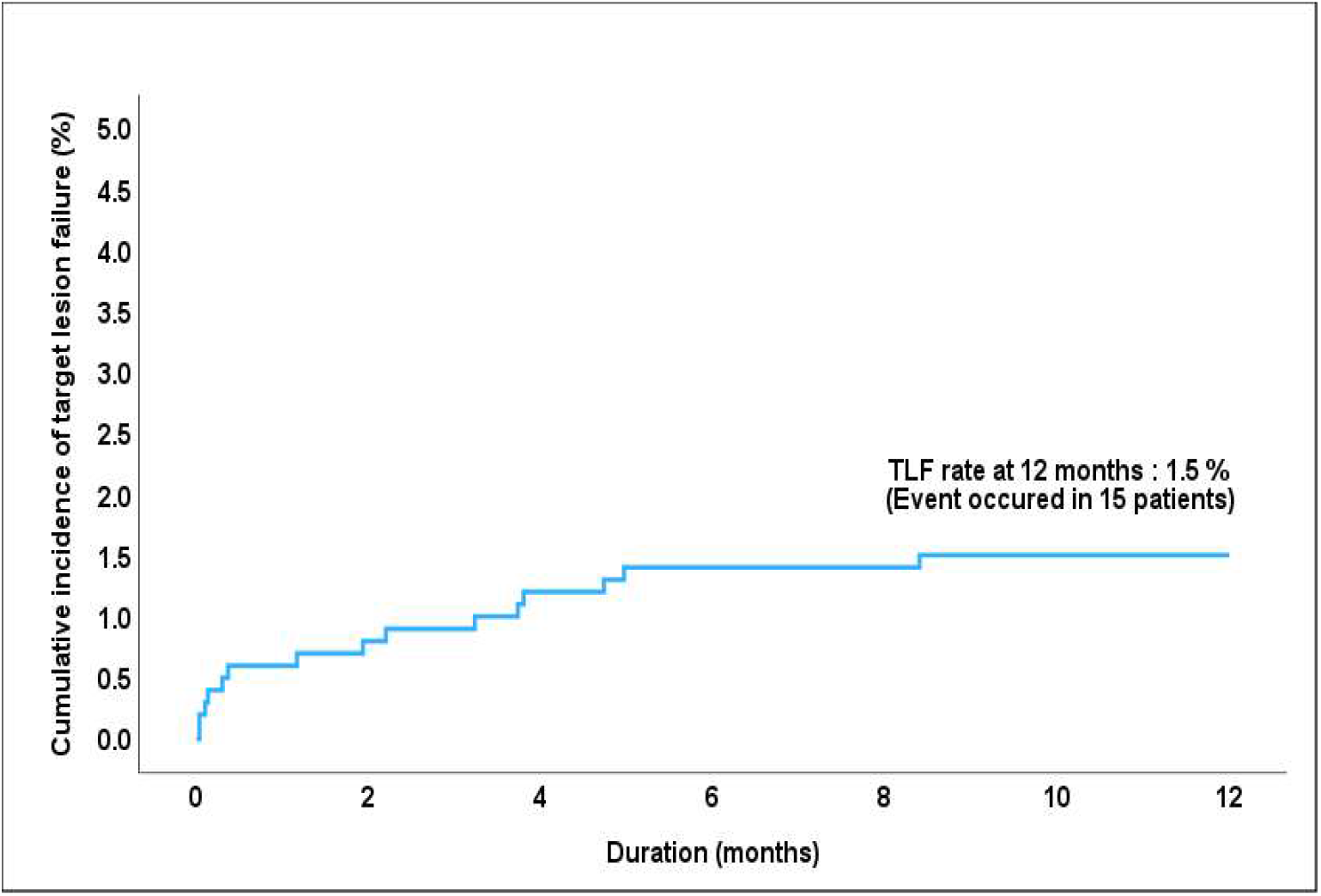
Cumulative incidence of the primary endpoint, target lesion failure

**Figure 2:**
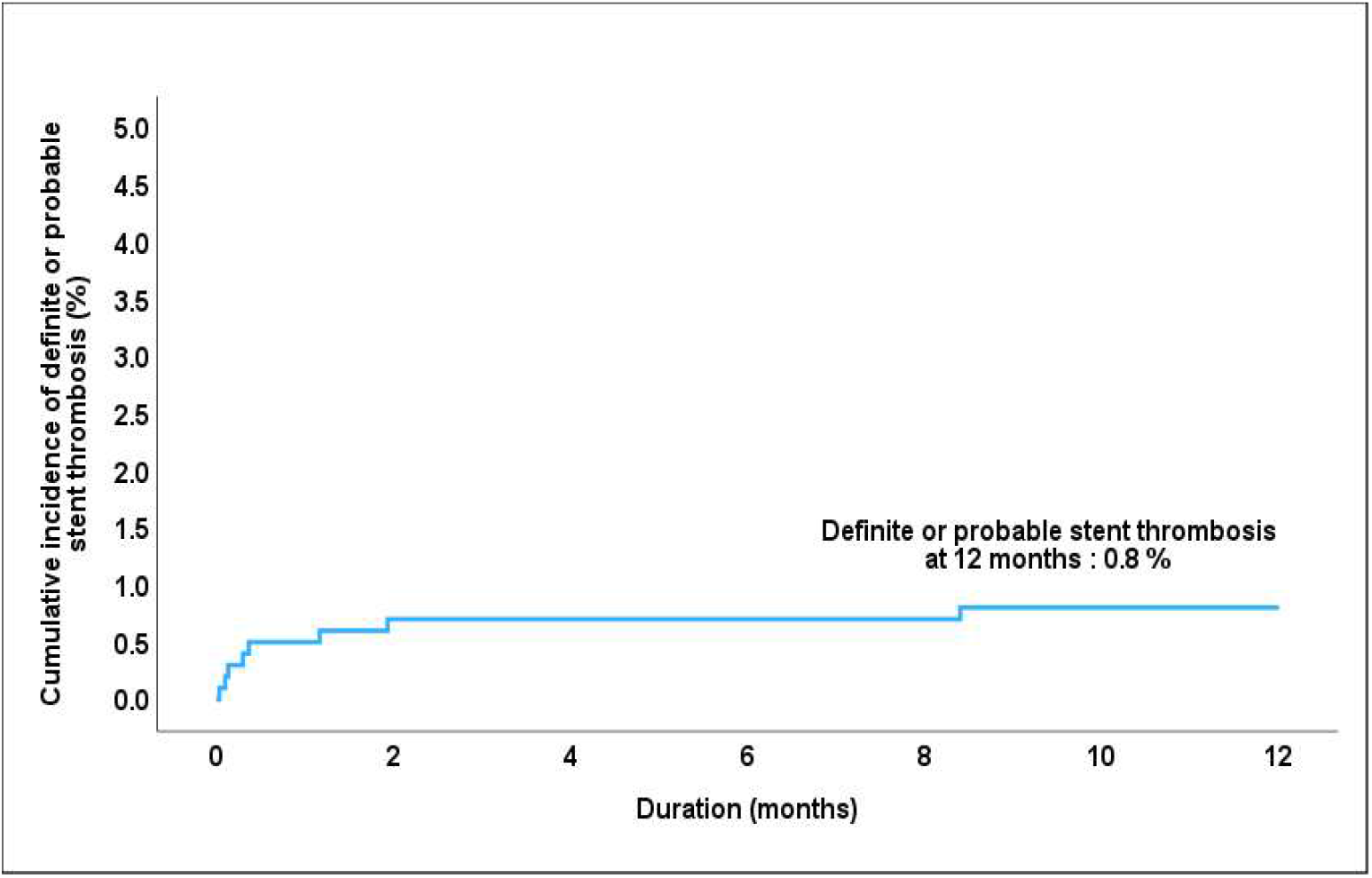
Cumulative incidence of the definite or probable stent thrombosis

## DISCUSSION

Continuous technological refinement of DES remains central to improving long-term outcomes following PCI. Although contemporary DES platforms have markedly reduced restenosis and stent thrombosis, persistent polymer exposure has been implicated in delayed arterial healing, chronic inflammation, and late adverse effects^9^. Consequently, two parallel design strategies have gained prominence: reduction in strut thickness and elimination of polymer coatings. Thin strut platforms reduce vessel injury and flow perturbation, while polymer-free technologies aim to mitigate chronic inflammatory responses associated with durable or slowly degrading polymers^5,6,10^ . Within this paradigm, everolimus represents a cornerstone antiproliferative agent owing to its favourable pharmacokinetic profile, potent inhibition of smooth muscle cell proliferation, and established clinical safety across multiple DES generations ^7^.

In this context, the present multicenter registry provides contemporary real-world evidence on the performance of a thin-strut polymer-free everolimus-eluting stent in a predominantly ACS population. At 12 months, rates of TLF and stent thrombosis were low, accompanied by high procedural and device success. The temporal distribution of events demonstrated early clustering within the first 6 months, followed by a plateau, suggesting early risk stabilisation and durable mid-term safety. These findings support the clinical reliability of the ISAR SUMMIT stent in complex and high-risk population.

These findings are consistent with evidence from randomised trials evaluating other polymer-free limus eluting stent platforms. In the ReCre8 trial, a non-inferiority trial enrolling 1502 all-comer patients, the polymer-free amphilimus-eluting stent demonstrated noninferiority to the permanent polymer zotarolimus-eluting stent, with 12-month TLF rates of 6.2% and 5.6% respectively^11^. Likewise, in the ISAR-TEST 5 trial, a randomised evaluation of 3002 patients with minimal exclusion criteria, a polymer-free sirolimus and probucol eluting stent was noninferior to the zotarolimus eluting stent at 12 months, with comparable rates of the primary composite endpoint (13.1% vs 13.5%) and definite or probable stent thrombosis (1.1% vs 1.2%)^12^. Consistent with these randomised data, a prospective all-comer cohort of 1664 patients comparing polymer-free stents with contemporary polymer-coated DES reported low and comparable 12-month TLR rates (1.7% vs 2.3%), with no significant differences in target vessel revascularisation or major adverse cardiac events^13^.

Evidence from contemporary meta-analyses further contextualises these findings. Updated systemic reviews comparing polymer-free DES with biodegradable or permanent polymer DES consistently demonstrate comparable rates of target lesion revascularisation, myocardial infarction, mortality, and stent thrombosis at short and mid-term follow-up^6,14^. While some device-specific signals have been reported, such as lower cardiac mortality with selected polymer-free platforms, these effects appear heterogenous and underscore the importance of evaluating individual device characteristics rather than polymer strategy alone. Overall, pooled data support polymer-free DES as a safe and effective alternative within modern PCI practice.

Collectively, these data suggest that polymer-free DES platforms achieve clinical outcomes that are at least non-inferior to best-in-class polymer-based DES. Importantly, this apparent equivalence should not be interpreted as a limitation of polymer-free technology. Rather, it reflects the substantial advancements achieved in contemporary DES design overall. As highlighted by prior investigators, focusing on polymer presence alone may oversimplify a multifactorial construct in which drug selection, elution kinetics, strut thickness, surface modification, and the scaffold architecture interact synergistically to determine vascular healing and long-term outcomes. Within this evidence framework, the 12-month TLF rate of 1.5% observed in the TRANSEVER registry, achieved in a predominantly ACS cohort with a high prevalence of diabetes and complex coronary disease, reflects the robust safety and efficacy profile of the polymer-free everolimus-eluting ISAR SUMMIT stent in contemporary real-world practice.

Supportive mechanistic evidence for polymer-free everolimus-eluting platforms is provided by first-in-human studies employing advanced surface modification techniques. In the early clinical evaluation of a titanium dioxide-coated polymer-free everolimus eluting stent, excellent angiographic and optical coherence tomography outcomes were observed, including low late lumen loss and complete strut coverage at mid-term follow-up, without safety concerns^15^. These findings reinforce the biological plausibility that polymer-free delivery of everolimus, when coupled with biocompatible surface engineering and thin strut architecture, can promote rapid endothelialisation while maintaining antirestenotic efficacy.

Several limitations merit consideration. As an observational study without a randomised comparator, unmeasured confounding cannot be excluded. The absence of systemic intracoronary imaging limits mechanistic insights into vascular healing. In addition, extended long-term follow-up data remain unavailable and longer-term outcome data will be required to establish the sustained efficacy and safety of this stent.

## CONCLUSION

In this large, contemporary real-world population with a very high proportion of patients presenting with ACS, the polymer-free everolimus-eluting ISAR SUMMIT stent demonstrated favourable 12-month clinical outcomes, with low rates of target lesion failure and stent thrombosis. These results suggest that this novel device is both safe and effective for routine clinical use.

## Data Availability

The data supporting the findings of the study are available from the corresponding author upon reasonable request.

## Acknowledgements

This investigator-initiated study was supported by an unconditional educational grant from Translumina Therapeutics Private Limited. Study funds were administered through Clicebo Solutions Pvt. Ltd., an independent Contract Research Organization; the sponsor had no involvement beyond providing this grant. The authors gratefully acknowledge Dr. Priyadarshini Armabam for her contributions to data management and statistical analysis.

